# A Bioinformatics Pipeline for Estimating Mitochondria DNA Copy Number and Heteroplasmy Levels from Whole Genome Sequencing Data

**DOI:** 10.1101/2021.12.28.21268452

**Authors:** Stephanie L Battle, Daniela Puiu, TOPMed mtDNA Working Group, Eric Boerwinkle, Kent D Taylor, Jerome I Rotter, Stephan S Rich, Megan L Grove, Nathan Pankratz, Jessica L Fetterman, Chunyu Liu, Dan E Arking

## Abstract

Mitochondrial diseases are a heterogeneous group of disorders that can be caused by mutations in the nuclear or mitochondrial genome. Mitochondrial DNA variants may exist in a state of heteroplasmy, where a percentage of DNA molecules harbor a variant, or homoplasmy, where all DNA molecules have a variant. The relative quantity of mtDNA in a cell, or copy number (mtDNA-CN), is associated with mitochondrial function, human disease, and mortality. To facilitate accurate identification of heteroplasmy and quantify mtDNA-CN, we built a bioinformatics pipeline that takes whole genome sequencing data and outputs mitochondrial variants, and mtDNA-CN. We incorporate variant annotations to facilitate determination of variant significance. Our pipeline yields uniform coverage by remapping to a circularized chrM and recovering reads falsely mapped to nuclear-encoded mitochondrial sequences. Notably, we construct a consensus chrM sequence for each sample and recall heteroplasmy against the sample’s unique mitochondrial genome. We observe an approximately 3-fold increased association with age for heteroplasmic variants in non-homopolymer regions and, are better able to capture genetic variation in the D-loop of chrM compared to existing software. Our bioinformatics pipeline more accurately captures features of mitochondrial genetics than existing pipelines that are important in understanding how mitochondrial dysfunction contributes to disease.

## 1 Introduction

Approximately 1 in 5,000 adults are diagnosed with a mitochondrial disease caused by a mitochondrial DNA (mtDNA) mutation[1]. Mitochondrial diseases are heterogeneous in their clinical manifestation and typically affect multiple organ systems[2]. For example, Leigh syndrome, the most common childhood mitochondrial disease, can be caused by more than 75 different mutations in nuclear or mitochondrial genes[3]. Some of the features include neurological symptoms, hypertrichosis, and dysmorphic features[2, 3]. MELAS, or Mitochondrial Encephalopathy, Lactic acidosis, and Stroke-like episodes, is a mitochondrial disorder where 80% of cases are caused by a mutation in a mitochondrial tRNA gene[4]. Sequencing patient DNA is commonly included as part of the diagnosis of mitochondrial diseases; therefore, being able to assess multiple features of mitochondrial genetics from genome sequencing data will be of significant benefit to human health.

The mitochondrion is a ubiquitous organelle with complex genetics. Unlike the nuclear genome, which is only present in two copies, there can be ∼1,000 to 10,000 copies of mtDNA in a cell[5]. The relative amount, or copy number, of mtDNA is associated with aging and overall-mortality[6, 7]. Additionally, the mitochondrial genome has a 17-fold higher mutation rate than the nuclear genome[8]. Thus, the mtDNA can exist in a state of heteroplasmy, where there is variation in the sequence of the different mtDNA molecules within a cell, or homoplasmy, where all mtDNA share the same sequence. Pathogenic mutations in mtDNA are usually present in a heteroplasmic state, and the level of heteroplasmies is directly linked to mitochondria function[2]. Both the quantity (as measured by copy number) and quality (as measured by amount of heteroplasmy) of mtDNA have been linked to disease[2, 7].

There are several software packages designed to take whole genome sequencing (WGS) data and extract mtDNA for variant identification. MToolbox[9] can extract mitochondrial reads from WGS or whole exome sequencing (WES) data to identify heteroplasmic single nucleotide variants (SNVs), insertions/deletions (INDELs), and haplogroup information. mtDNA-Server[10] which uses the program Mutserve, identifies heteroplasmy and works very well on large datasets. MitoAnalyzer[11, 12] performs both heteroplasmy calling and copy number calculations. Mity[13] is another software that detects heteroplasmy SNVs and INDELs from WGS data. These software attempt to address two basic features of mitochondrial genetics, sequence variation and copy number, and each has its own unique limitations. None of them attempts to recover sequencing reads at regions of low coverage, which is important for thorough variant discovery.

Here we present a bioinformatic pipeline to estimate mitochondrial copy number (mtDNA-CN) and heteroplasmy from WGS samples. The pipeline is able to obtain uniform coverage across chrM and remove contaminating nuclear-integrated mitochondrial sequences (NUMTs). Our pipeline also constructs a reference sequence for each sample and recalculates heteroplasmy variant frequency for each sample using its own reference. We optimized our pipeline to run efficiently on 10,000s of samples and have added sample contamination checks. The pipeline additionally annotates SNVs and INDELs to allow for better identification of true variation from sequencing or mapping errors.

## 2 Materials and Methods

### 2.1 Datasets

Datasets are from the Trans-Omics for Precision Medicine (TOPMed) program [14]. TOPMed studies provide WGS data at 30x genomic coverage using Illumina based next-generation sequencing technology. TOPMed WGS data must pass specific quality control metrics before it is released for use by the scientific community. Additional information on TOPMed WGS data generation and processing can be found here: https://www.nhlbiwgs.org/data-sets

We analyzed WGS data from the Atherosclerosis Risk in Communities (ARIC) study [15] and the Multi-Ethnic Study of Atherosclerosis (MESA) study [16]. Both ARIC and MESA are population-based longitudinal cohort studies with 3930 and 5370 WGS samples available, respectively. One sample in ARIC was excluded due to lack of proper consent. ARIC WGS samples were comprised of deep vein thrombosis and early-onset atrial fibrillation cases (< 10% of dataset) and controls. In the ARIC study, DNA for WGS were isolated from buffy coat using the Gentra Puregene Blood Kit (Qiagen), The ARIC cohort is 52% female, age range 45-74 at time of DNA isolation with the following racial backgrounds: 93% European American and 7% African American. MESA participants were required to have no known clinical CVD upon recruitment. In MESA, DNA was isolated from peripheral leukocytes using the Gentra Puregene Blood Kit. The MESA cohort is 53% female, age range 45-84 with the following racial backgrounds: 38% European American, 28% African American, 22% Hispanic and 12% Chinese American ancestry.

### 2.2 TOPMed Google Cloud Data Access and Extracting Metadata

The TOPMed datasets used for our study were accessed using Google Computing Services (Supplementary Figure 1). Samples were processed in batches of 30. We mounted the CRAM and CRAI files using fusera, extracted chrM/NUMT reads using “samtools view –T hg38-reference-file –F chrM chr1:629084-634422 chr17:22521366-22521502” and generated output BAM files.

### 2.3 Processing from FASTQ files

The pipeline is designed to run on aligned human WGS data (Figure 1). Prior to running the pipeline, FASTQ files should be trimmed and aligned to the whole human genome assembly using an aligner which generates SAM/BAM/CRAM output alignments. SAM format alignment files can be convert to BAM/CRAM format using Samtools software. The alignment files must be sorted (samtools sort) and indexed (samtools index). The total number of reads and aligned reads counts (samtools view) are used for mtDNA-CN estimation.

**Figure 1:**
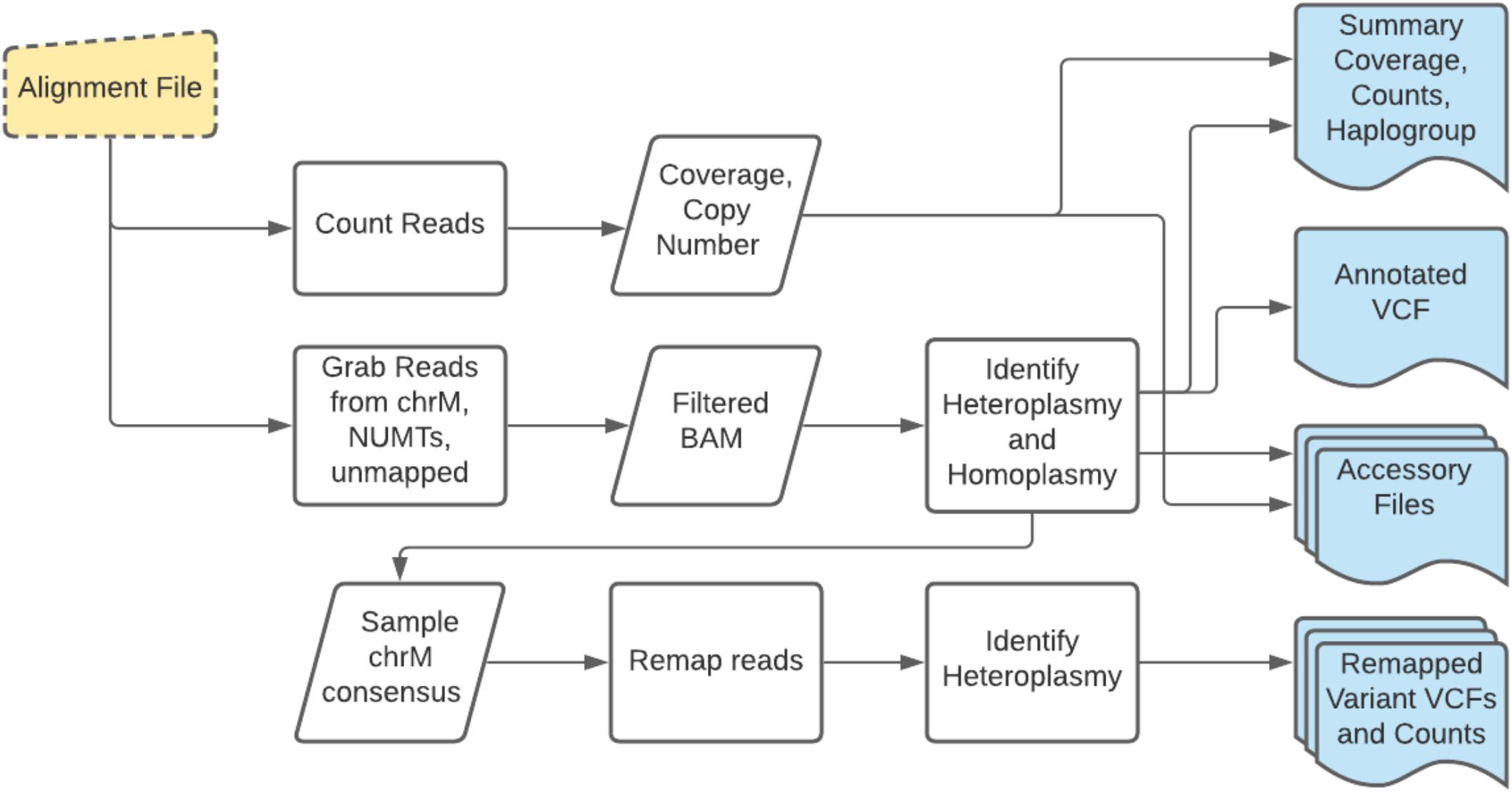
Overview of mtDNA-CN and Heteroplasmy Analysis Pipeline

### 2.4 Calculations for mtDNA-CN

The general calculation for mtDNA-CN is shown below: 2 x (chrM coverage) / (genome coverage)

Mitochondrial genome coverage is calculated as the number of mapped reads to chrM times the read length divided by the length of chrM. Genome coverage is calculated using three different methods described below:

1. coverage calculated by TOPMed from mappable reads passing data quality filters aligned to a sex specific mappable genome
2. (mapped bases) / (3,217,346,917 bases in human genome)
3. (mapped bases) / (3,160,119,502 bases if female) or (3,110,712,762 bases if male) The comparison of these different copy number metrics is described in the Results section.

### 2.5 Extracting and Re-Mapping Reads to a Circularized chrM

The WGS data was mapped to the human genome build GRCh38 (Figure 2). We used Samtools v1.11 to extract reads that mapped to chrM and the NUMT regions (hg38 chr1:629084-634422 and chr17:22521366-22521502; hg19 chr1:564465-569708 chr17:22020692-22020827). We also retrieved unmapped reads where one mate mapped to chrM. We remapped the reads to a circularized version of chrM with position mt16569 extended downstream 300 bases and the chr1 and chr17 NUMT regions (Figure 2). This extended end was included in the ref.fa file which was indexed using “bwa index”. The reads were trimmed using “fastp”[17] and remapped using BWA v 0.7.17 with the following parameters: -p -v 1 -t 1 -Y -R “headerline” -v 1. Duplicate reads were removed using “samblaster --removeDups –addMateTags”. Alignments that spanned the chrM start-stop were split and kept as 2 alignments.

**Figure 2:**
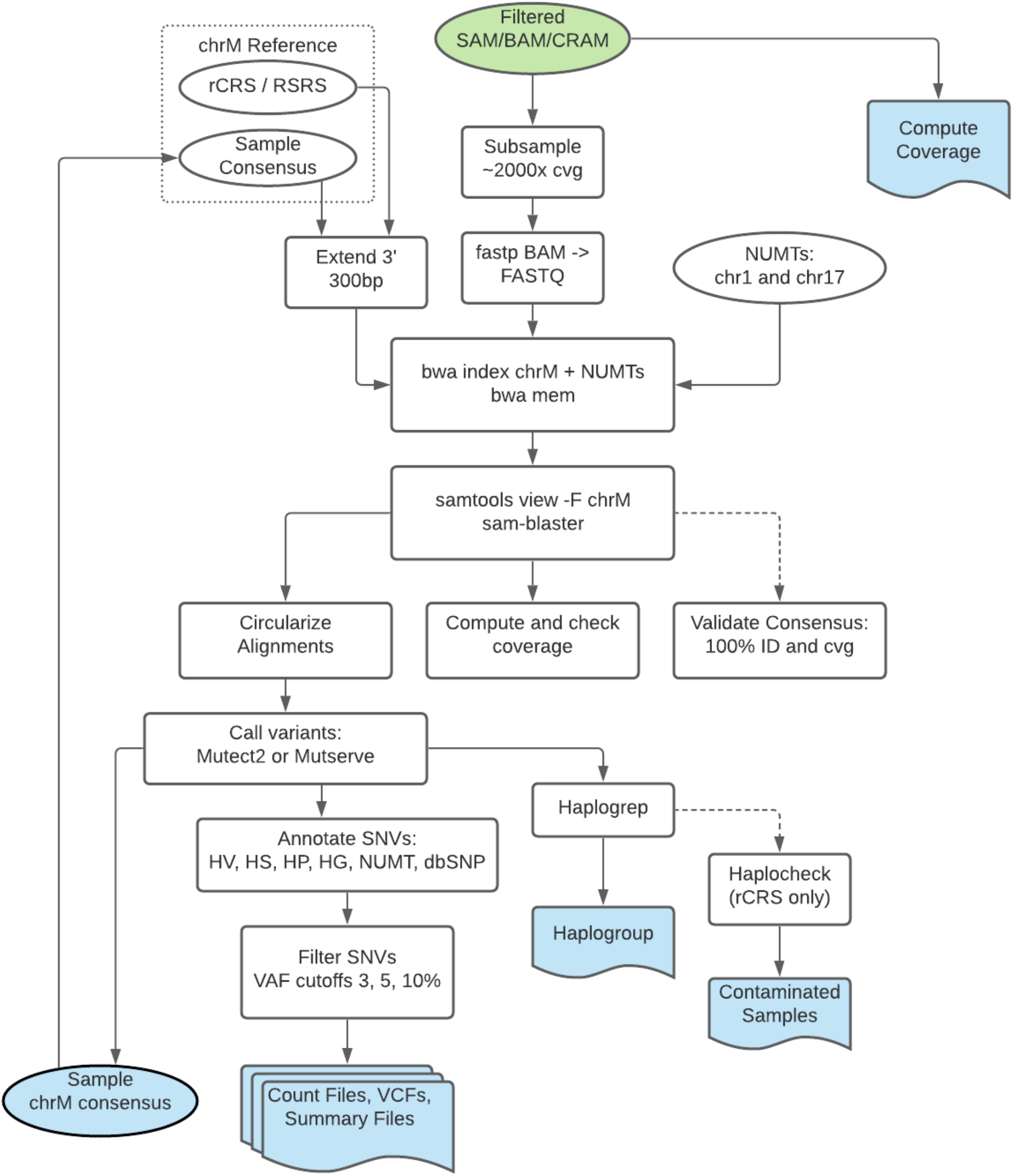
Detailed flowchart of pipeline processes. Input is BAM file (green) at top of diagram. Outputs are shaded blue.

### 2.6 Detecting Sample Heteroplasmy, Homoplasmy and Haplogroup

Prior to running the mtDNA heteroplasmy calling software, we downsampled the chrM reads for each sample to ∼2000x coverage (Figure 2). This increases the speed of our analysis pipeline while still retaining high enough coverage to have confidence in low level (3% variant allele frequency) heteroplasmy calls. We incorporated two programs for calling mtDNA heteroplamsy and homoplasmy, GATK Mutect2[18, 19] and Mutserve[10] (Figure 2). For both programs we used multiple pre-specified heteroplasmy VAF thresholds (3%, 5%, and 10%). We used the rCRS as the reference genome, although the RSRS is included as an optional reference genome. Mutect2 was run using default parameters and the output VCF was run through FilterMutectCalls command with the following additional parameters: *--min-reads-per-strand 2*. Mutserve was run using the following additional parameters: *--deletions --insertions --level 0*.*01*.

### 2.7 Variant Annotation

Variant annotations are included in the VCF output files. In addition to annotations created by Mutect2, we included annotations that add genomic and biological context to the variant sites identified. The following annotation files are provided on GitHub: 1) mitochondrial hypervariable (HV) regions; 2) chrM homopolymer (HP) regions defined as 5 or more Cs in a row, 1 mismatch, +/-1 bp on the ends; 3) hotspot regions (HS) known to be highly variable; 4) coding regions (CDS); 5) manually curated haplogroup specific SNV’s (HG) which contains SNVs which occur in in 80% of the cases in a particular haplogroup; 6) NUMT SNV’s identified by aligning the 2 NUMTs (chr1,chr17) to rCRS using MUMMer nucmer and show-snps; 7) dbSNP are variants identified in dbSNP database[20]. We also annotate variants with their Combined Annotation Dependent Depletion (CADD) score (version 1.3), which scores the functional consequence of mitochondrial variants [21].

### 2.8 Generate Consensus Sequence and Validate Alignments

We use “bcftools consensus” to generate a new mitochondrial consensus fasta sequence for each sample incorporating homoplasmies and major alleles from mutect2 output. This sequence was circularized (300bp from position mt1 added after position mt16569) and indexed using “bwa index”. The 2000x coverage reads were aligned using “bwa mem”. Exact alignments (100% identity, 100% length) were converted to BED format and merged using “bedtools merge –d -5”, making sure the reference was fully covered.

### 2.9 Contamination Check

Due to the exclusive maternal inheritance, each sample should have only 1 dominant haplogroup detected. We ran haplocheck[22] on samples after haplogroup identification. Haplocheck output file lists all samples and contamination status. We removed samples marked as contaminated with a contamination level of 3% or more from downstream analyses. We used a database of 1098 haplogroup specific SNVs, which contains SNVs that occur in 80% of the samples in a particular haplogroup[23]. We also created a database of 217 of NUMT specific SNVs based on the alignment of hs38DH chr1 and chr17 NUMTs to rCRS. Variants with the “NUMT” annotation have an alternate allele that resembles a nuclear sequence at chr1 or chr17 but at chrM coverage levels. The number of samples with HG SNPs belonging to multiple haplogroups would was very low (∼1 in 1000) and these samples were visually inspected.

### 2.10 Statistical Analyses

Statistical analyses were performed using R version 4.0.4. To test for an association with age and mtDNA-CN, we first adjusted heteroplasmy for covariates and then took the scaled residuals to run the association. We adjusted binary coded heteroplasmy (where ‘0’ means no heteroplasmic sites and ‘1’ means at least one heteroplasmic site) using a binomial generalized linear model. We included the following covariates: age, sex, race, and collection center. A linear model was used to run the association with scaled residuals and age or mtDNA-CN. Average heteroplasmy count was performed in R after removing outlier counts that were 3 or more standard deviations away from the mean.

## 3 Results

### 3.1 Overview of Pipeline

Our goal was to create a bioinformatics pipeline that incorporates multiple features of mitochondrial genetics to readily facilitate downstream analyses. This pipeline includes four main parts: 1) calculation of mtDNA-CN; 2) identification of heteroplasmic and homoplasmic variants (referred to as “first iteration”); 3) generation of the chrM consensus sequence (incorporating homoplasmies and major heteroplasmic alleles in sequence, using the rCRS as the backbone) for each sample (referred to as “second iteration”); and 4) re-calling heteroplasmy for each sample mapped against its own consensus sequence. Our mitochondrial genetics pipeline produces three main outputs. The first output is a mtDNA-CN summary file with read count, coverage, and mtDNA-CN counts. The second output is a variant summary information file, which includes haplogroup, a count of mtDNA homoplasmic sites, heteroplasmic sites, SNVs, INDELS at all locations and at non-homopolymer regions. The third output consists of VCF files of the annotated heteroplasmic and homoplasmic sites. Re-calling heteroplasmy against a sample’s own reference generates the additional summary and VCF files. The code is available on GitHub: https://github.com/danarking1/HP.

### 3.2 Computational Speed

We utilized Google Cloud to filter the chrM and nuclear integrated mitochondrial sequence, or NUMT, reads from TOPMed samples. The sample alignment files were processed in batches of 30 on a single processor and took ∼2 minutes per sample to complete. Running in parallel with a maximum 240 jobs at one time, it took ∼1.5 days to process the 90K samples. When it comes to computational speed, our pipeline is designed to handle large genomics dataset of 10,000s of samples quickly and efficiently.

### 3.3 Recovering Low Coverage Areas

Accurately aligning reads to the chrM can prove challenging. First, chrM is circular, and commonly used aligners expect linear chromosomes. Also, chrM reads can falsely align to NUMTs in the nuclear genome. From the provided TOPMed metadata, we first checked the uniformity of coverage across chrM (Figure 3, red line). As expected, we saw noticeable dips at the ends of the chrM and at other sites known to have low coverage. Position chrM:310 and chrM:460 lie within polycytosine tracts (chrM:300-320 AAACCCCCCCTCCCCCGCTTC and chrM:450-470 TATTTTCCCCTCCCACTCCCA) and have previously been reported to have low coverage in sequencing data[24]. Low coverage at three mitochondrial hypervariable regions (HVRs) due to polycytosine tracts in these regions have also previously been reported[25]. This is due to polymerase slippage at regions of low nucleotide complexity either during sequencing, library PCR, or within the cell during chrM replication[24].

**Figure 3:**
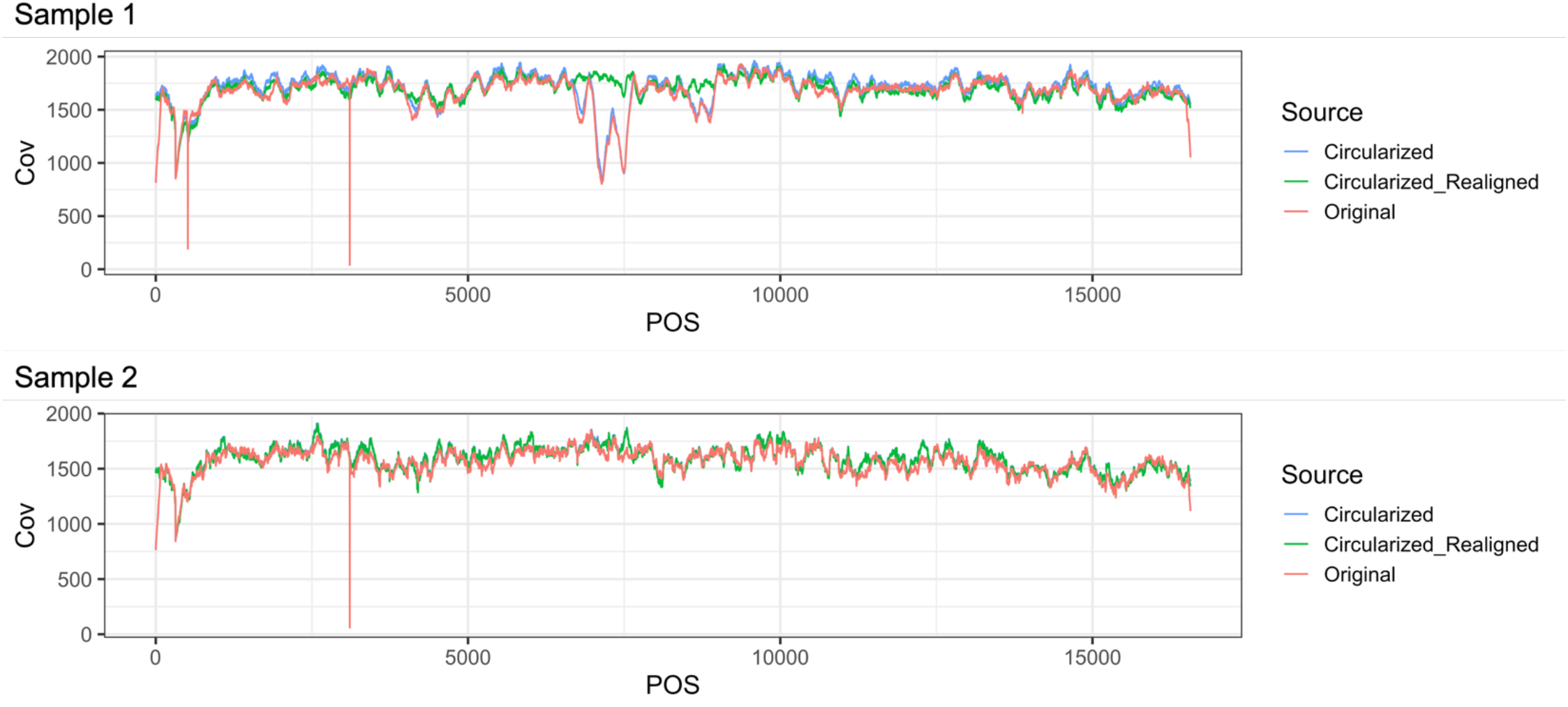
Coverage Correction Coverage for two samples in ARIC plotted across the full length chrM. Sample 1 belongs to haplogroup L1 and sample 2 belongs to haplogroup H. Reads were deduplicated prior to plotting. The red line is chrM coverage from the original alignment file. The blue line is the chrM coverage after aligning the reads to the circularized reference. The green line is the chrM coverage after realigning the reads to the sample’s reconstructed chrM reference sequence. All reads were downsampled evenly for plotting.

To recover reads at low coverage regions we started by realigning the reads to the circularized version of chrM. Due to the “edge” effect (low coverage at the start/stop of chrM), and the similarity of the chr17 NUMT to chrM D loop (bwa mem minimum alignment score of 30), the coverage in the D loop region is about ∼40% that of the chrM median coverage. By aligning to the circularized MT, the average D loop coverage increased to more than ∼90% of the average total chrM coverage (Figure 3, blue line). Realigning reads to a sample’s unique consensus chrM sequence, increased the coverage of other low coverage regions. For example, in Figure 3, the sample 1 from ARIC had a major dip in coverage upstream of position 7500, which was only recovered when using the sample’s unique consensus as a reference for alignment (Figure 3, green line).

Sample 2 had a more subtle difference in coverage but also showed a notable increase in coverage at the start/stop of chrM. Our pipeline prioritizes uniform chrM read coverage prior to heteroplasmy calling; however, it is still possible for some samples to have low average depth of coverage. It is important to inspect the coverage depth of outlier samples and variants. However, with our method of realigning to a sample’s unique reference, we are able to attenuate large differences in coverage across chrM.

### 3.4 Contamination Check

We included the program haplocheck[22] in our pipeline to output the contamination status of samples. Haplocheck uses the haplogroup of a sample and evaluates the reads for the presence of variants from another haplogroup, which would suggest contamination in the WGS sample. Four samples out of 3929 in ARIC and 10 samples of out 5370 in MESA had a haplocheck contamination level of 3% or more. For our purposes, the potentially contaminated samples were excluded from our downstream analyses. Although depending on the biology of the samples in question, it may be worth investigating the “contaminated” samples further. The inclusion of additional sample QC checks, like haplocheck, in our pipeline allows for the user to easily identify samples of poor quality, as their results could confound downstream analyses.

### 3.5 mtDNA-CN calculation comparisons

Mitochondrial DNA copy number is a metric commonly used for mitochondrial quantity in a cell or tissue and is associated with mitochondrial function[26]. It is based on the ratio of mtDNA to nuclear DNA. To identify the best method to calculate mtDNA-CN, we used the same basic equation defined as two time the ratio of chrM coverage to genome coverage. The chrM coverage was relatively uniform across all haplogroups in ARIC and MESA cohorts (Supplementary Figure 2). We calculated the genome coverage using three different approaches to determine whether the subtle differences in the mtDNA-CN calculation would have an impact (See Methods). For method 1 we used the average genome coverage included in the TOPMed metadata file. For method 2, we “recomputed” the average genome coverage based on the number of bases sequenced divided by the standard human genome size. Method 3 is the sex-adjusted genome coverage, which is the total number of bases divided by the genome size for females or males. Due to the sex chromosomes, the female genome is 1.02x larger than males. Additionally, TOPMed provides an mtDNA-CN metric computed using the program fastMitoCalc[12] and is available for download for TOPMed datasets.

We observed a high correlation between the different mtDNA-CN metrics (all r>0.98, Supplementary Figure 3) and the overall distribution of the mtDNA-CN values were similar for all four CN metrics (Supplementary Figure 2). Given their similarity, we arbitrarily checked one mtDNA-CN metric, sex-adjusted metric, for any haplotype bias and found no major differences in mtDNA-CN across all haplogroups (Supplementary Figure 2). Although we did note that the overall mtDNA-CN metric is higher in the ARIC cohort compared to MESA, which may be due to a difference in the DNA source.

We next sought to understand how each mtDNA-CN metric performed in an association with known associated phenotypes (Figure 4). We and others have previously shown that mtDNA-CN measured from peripheral blood decreases with age and is higher in females than in males[7, 27]. The mtDNA-CN associations were tested using a liner regression model, adjusted for age or sex, race, and collection center. As expected, samples from older individuals had a lower mtDNA-CN and females had higher mtDNA-CN than males. For additional assessment of the mtDNA-CN metrics, we also investigated the association of mtDNA-CN with a copy number polygenic risk score (PRS) in the ARIC cohort. PRS was calculated from SNPs identified from GWAS performed in 465,809 individuals in the UK Biobank and Cohorts for Heart and Aging Research in Genomic Epidemiology (CHARGE)[28]. When including age, sex, race and center as covariates in our linear regression analysis, all 4 mtDNA-CN metrics were associated with the mtDNA-CN PRS, with the TOPMed mtDNA-CN provided metric having greatest significance (Figure 4).

**Figure 4:**
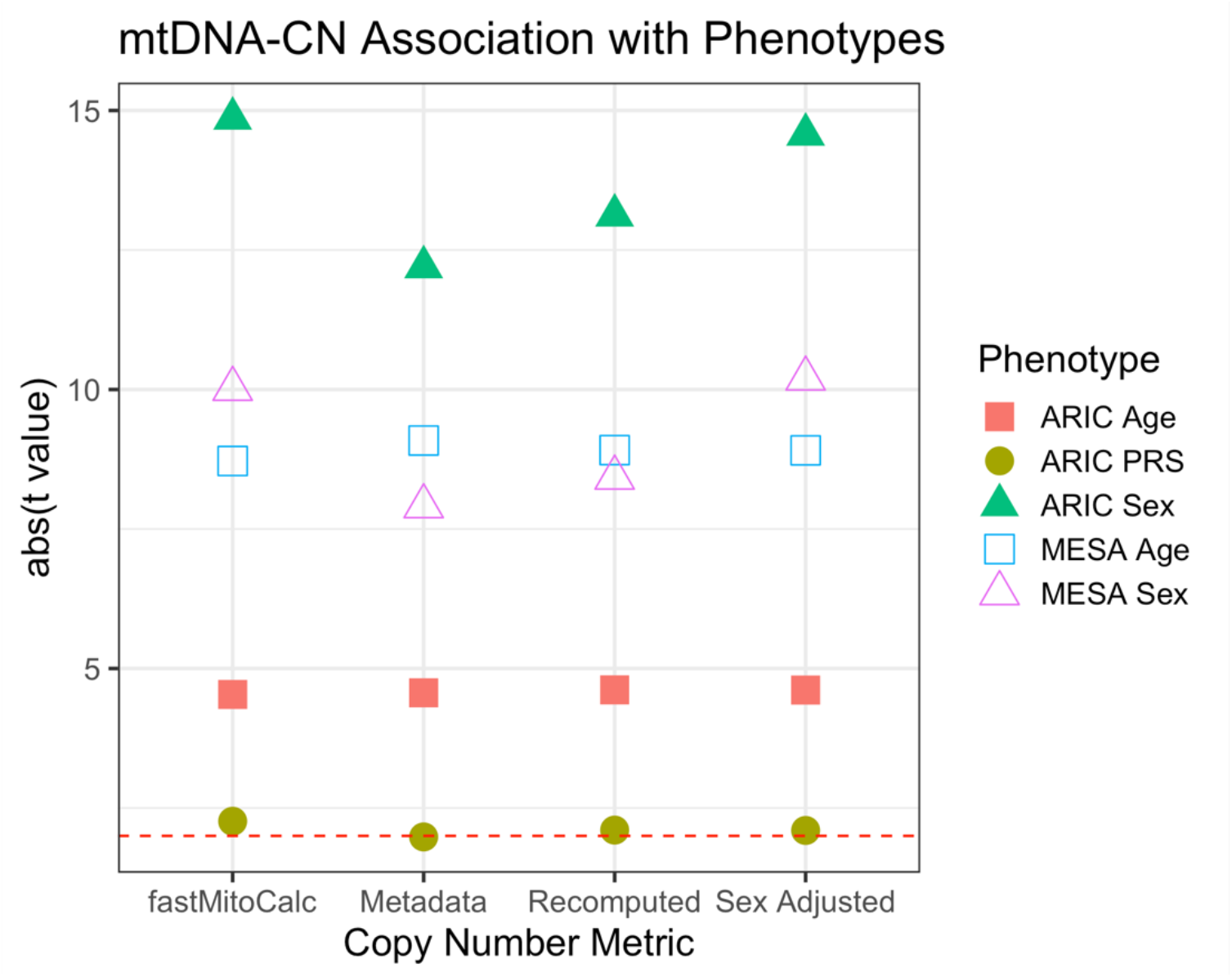
Age and Sex Association with mtDNA-CN. Linear regression results for mtDNA-CN and age or sex in ARIC (n = 3608, left) and MESA (n = 4579, right) cohorts. Polygenic Risk Score (PRS) are only in ARIC (n = 3018). Red dashed line indicates t value of 2.

To identify the best metric for mtDNA-CN, we ranked each metric based on the strength of the association with known phenotypes as measured by p values (Supplementary Figure 4). We used the Kendall’s W test to determine if there was an agreement among the rankings. We observed that there is no significant agreement in ranking, indicating that no mtDNA-CN metric is significantly better than the others. Our pipeline by default will calculate mtDNA-CN as the total number of bases mapped divided by a standard reference genome size (3.22G), referred to as “recomputed” within this manuscript. In our datasets the difference in effect size between the mtDNA-CN calculations and their association with known phenotypes was subtle, suggesting that the mtDNA-CN metric from WGS is generally robust and the specific calculation should not have a major effect on downstream analyses.

### 3.6 Characterizing Heteroplasmic Variants

Mitochondrial SNV heteroplasmies identified by next generation sequencing can be reliably detected at 3% variant allele frequency (VAF) with 1000x chrM coverage[29] and thus, we performed our analyses on heteroplasmy SNVs called as low as 3% VAF. We recommend down-sampling to a higher average coverage if identifying heteroplasmic variants below 3% and being mindful of sample quality via mtDNA-CN. At 3% VAF, the average heteroplasmic SNV site count was 0.9 and 1.2 for ARIC and MESA, respectively, excluding outlier counts that were 3 or more standard deviations from the mean. We noticed that samples with heteroplasmic site count greater than 5 have significantly lower mtDNA-CN (p = 0.02 in ARIC and p < 2×10^−16^ in MESA). Extreme mtDNA-CN values may be an indicator of poor sample quality since mtDNA-CN is more affected by technical procedures such as DNA isolation techniques. Even though we did not observe a haplogroup bias with chrM coverage or mtDNA-CN, some haplogroups did have significantly different heteroplasmy counts (Supplementary Figure 8). The L haplogroups have a higher number of homoplasmic variants due to the rCRS reference being most similar to H haplogroups and least similar to L (Supplementary Figure 7).

Both Mutect2 and Mutserve had similar distributions of heteroplasmic site counts (Supplementary Figure 6); however, Mutect2 detects more samples as having one or more heteroplasmic sites than Mutserve. For example, Mutect2 identified 2084 samples with one or more heteroplasmic sites in ARIC compared to 1833 by Mutserve. In both ARIC and MESA, the vast majority of variants were identified by both programs, with Mutect2 identifying over 3000 additional variants in ARIC and over 6000 additional variants in MESA (Figure 5A). Of variants that are uniquely identified by either program, only Mutect2 detected variants in chrM hypervariable regions (Figure 5A). When we plotted the VAFs for each SNV in each sample identified by Mutect2 and Mutserve, there were variants where one software called the position a homoplasmy and the other software called the variant a heteroplasmy. This was observed in both cohorts and these variants were almost exclusively in the mitochondrial D-loop (Figure 5B). As a result of the hypervariable regions and poly-C homopolymer tracts in the D-loop, many of the variants identified in this region had low base quality for the alternate allele or had other annotations suggestive of sequencing or technical errors. By performing a comparison of Mutect2 and Mutserve, we found that the genomic substructures of chrM plays a significant role in variant identification. Nevertheless, Mutect2 and Mutserve are largely similar in the variants they identify.

**Figure 5:**
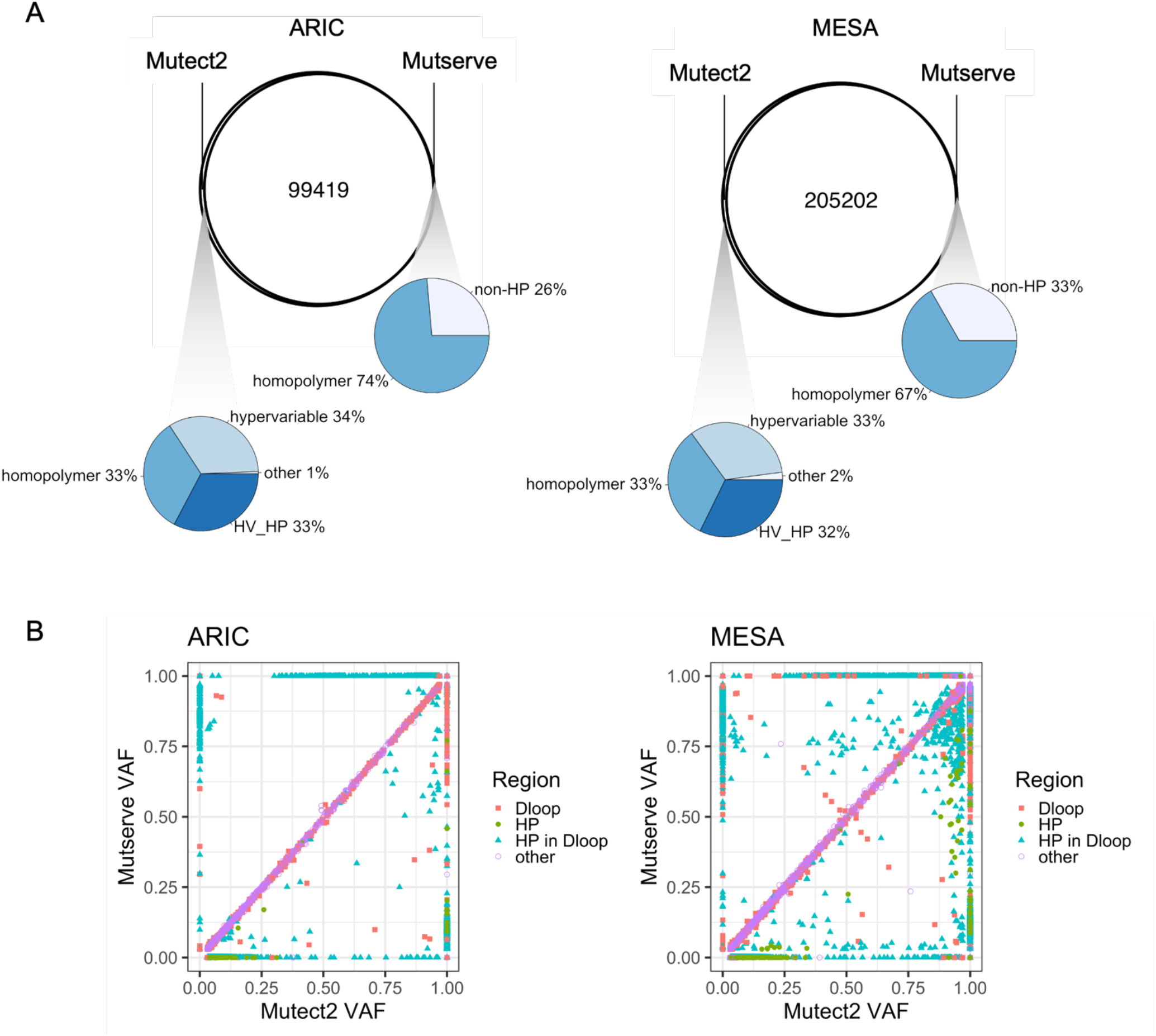
SNVs Called by Mutect2 and Mutserve Venn diagrams show the overlap/differences in the variants called by Mutect2 and Mutserve. The bottom panel shows scatterplots of VAFs of SNVs called at 3% VAF threshold by both software. Colors indicate the location of the variant, in the D-loop (red), in homopolymer (HP) regions (green), in a HP region within the D-loop (blue) or in another region (purple).

To cross-validate the variants we identified, we determined how many of the variants were also present in the gnomAD v3 database of over 56k WGS samples[30]. We took 12,749 unique chrM homoplasmic and heteroplasmic variants from gnomAD. Of the variants we identified, 99% of sites in ARIC and MESA are also in the gnomAD database. It should be noted that the gnomAD variants had a lower frequency cut-off of 10% instead of the 3% used in our study. These results support the conclusion that our heteroplasmy calling pipeline is likely identifying true variants within the general population, using a lower frequency threshold for defining heteroplasmic variants.

### 3.7 Detection of True versus False Positive Heteroplasmic Variants

Since we observed unique identify variants from the same datasets, we asked if one software was detecting more false positive heteroplasmic variants. To test this, we generated 30 simulated datasets representing 30 main haplogroups. Each data was simulated to have 150 bp paired-end reads with a random base pair error rate of 0.01. We introduced 43 heteroplasmic sites, 8 INDELs and 35 SNVs into each simulated sample dataset at an average of 18% VAF distribution (range 15% - 20%). The reads were all mapped using bwa to the rCRS reference. We compared the number of heteroplasmic sites identified by the first iteration Mutect2, Mutserve as run by our pipeline, the online based client Mitoverse, which uses Mutserve, and MToolBox. MToolBox is the only program that uses the aligner GSNAP [31]. We ran the 30 datasets through the different variant callers and counted the number of heteroplasmic sites identified (Table 1). Overall, Mutect2 had the fewest number of false-positives and false-negative calls for both the first and second iteration of variant calling. Mutserve detected more false positive heteroplasmic sites, suggesting that the uniquely identified heteroplasmic variants in Figure 5 were less likely to be real variants, particularly as they occur in homopolymer regions. Since Mutect2 performed the best on our simulated data, we used the Mutect2 variant calls for all analyses moving forward.

**Table 1:** Average Number of Heteroplasmic Sites identified across simulated data

|  | All Identified sites | True positives | False positives |
| --- | --- | --- | --- |
| 100x coverage |  |  |  |
| 1<sup>st</sup> iteration Mutect2 | 43.27 | 41.80 | 1.91 |
| Mutserve | 37.07 | 33.57 | 3.50 |
| Mitoverse | 38.00 | 34.30 | 3.70 |
| MToolbox | 50.93 | 41.90 | 9.03 |
| 2000x coverage |  |  |  |
| 1<sup>st</sup> iteration Mutect2 | 43.03 | 42.93 | 1.00 |
| Mutserve | 39.73 | 38.90 | 1.92 |
| Mitoverse | 37.80 | 36.83 | 2.07 |
| MToolbox | 52.33 | 42.97 | 9.37 |

### 3.8 Second Iteration Mutect2 – Calling Variants Against the Sample Reference

In its essence, homoplasmy represents inter-individual mitochondrial variation, while heteroplasmy represents intra-individual variation. To leverage this observation, we generate a unique reference for each sample using its own mtDNA consensus sequence generated from homoplasmic sites and major allele heteroplasmic sites called by Mutect2. Heteroplasmic variants are then identified by remapping reads against the sample’s unique mtDNA reference sequence. We refer to this as the “second iteration” of heteroplasmy calls. The SNV heteroplasmic site count decreased an average 0.3 counts in ARIC and 0.4 counts in MESA, suggesting a reduction in false-positive heteroplasmies. Notably, some haplogroups had significantly different heteroplasmic site counts (Supplementary Figure 8)

We expect the vast majority of the SNVs identified by the first iteration of heteroplasmy calling to also be identified by the second iteration heteroplasmy calling. Surprisingly, we observed a substantial number of SNVs that, at first glance, appeared to be unique to the either iteration (Figure 6). However, the majority of variants that appear to be uniquely identified are simply the result of an allele switch between the reference and alternate allele. For example, ARIC sample SRR8015883 has a reference A allele at position 57 and a heteroplasmic G at 9% VAF identified in the second iteration of variant calls. In the first iteration position 57 of the rCRS ref allele is T and the A allele is recorded as 91% VAF and the G allele is measured at 9% VAF. The T allele is recorded in that sample’s reference sequence in the second iteration and thus, the only variant outputted at position 57 is the A allele.

**Figure 6:**
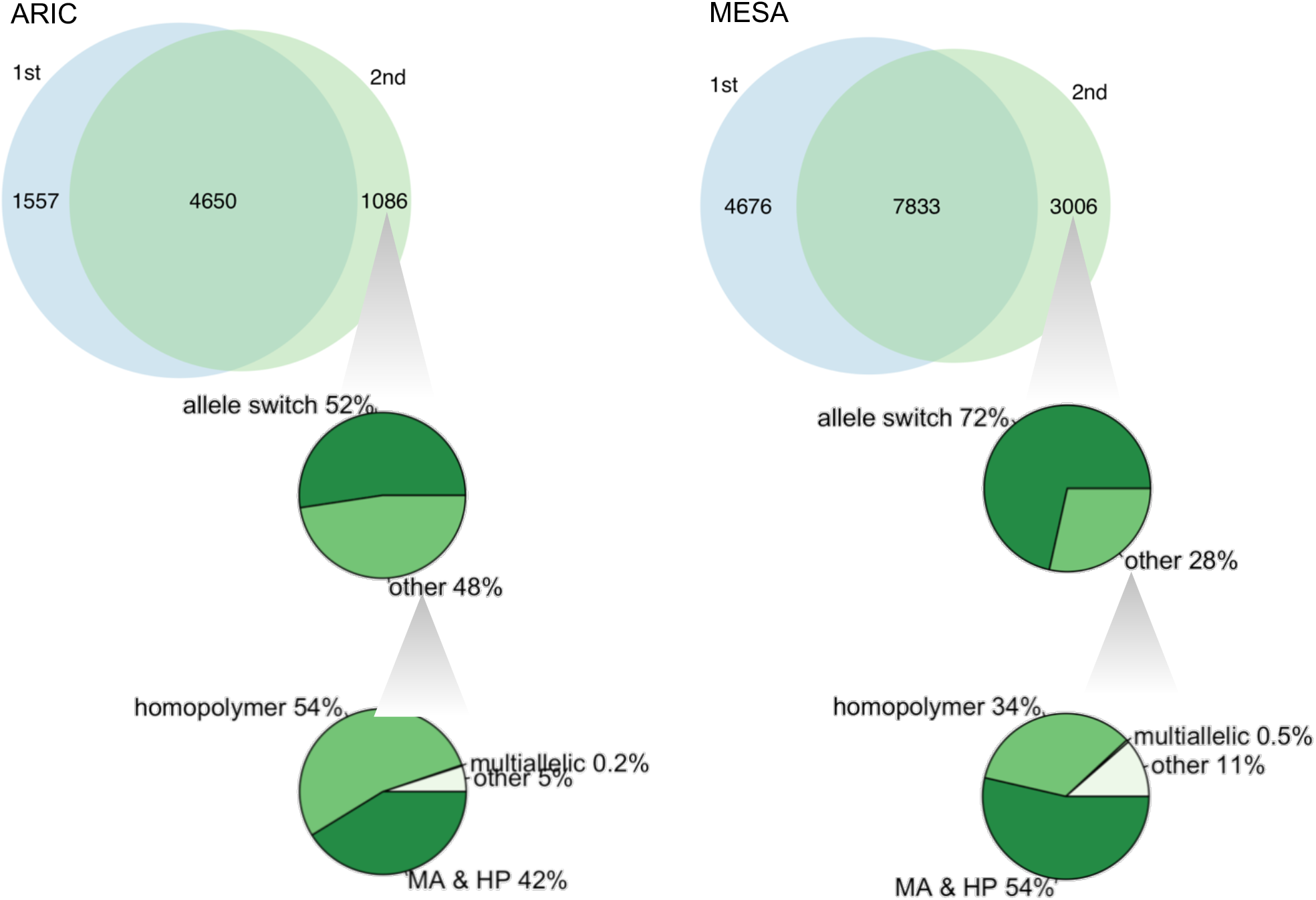
Comparison of heteroplasmic and homoplasmic SNV site counts between 1^st^ and 2^nd^ iteration Mutect2 variant calling (top panel). The first level of pie charts show the percentage of variants that “appear” different due to a ref/alt allele switch. The second level of pie charts show the percentage of non-allele switch (or “other”) variants that were multiallelic, in a homopolymer region, both (MA & HP), or neither (other).

We further investigated the SNVs that differed between the first and second iteration of heteroplasmy calling that were not due to a simple ref/alt allele change. Of these differential SNVs, 95% in ARIC and 88% in MESA are within a homopolymer region (Figure 6). The majority of first iteration unique sites were at positions 302, 16183, and 310 while the majority of second iteration unique sites were at positions 310 and 16182. These sites were within the homopolymer regions at positions 296-318 and 16178-16193. We also found that 42% and 54% respectively were also multiallelic sites. For example, ARIC sample SRR8015858 had a SNV A > C (3% frequency) at position 302 identified by the second iteration heteroplasmy calling. This sample had two INDELs identified at this same position, AC > A (5%) and AC > ACC (14%). For this sample, in the first iteration, the variants at position 302 were three INDELs, A > AC (6%), A > ACC (74%), A > ACCC (15%). Homopolymer regions make variant calling complicated and within our pipeline we found that these sites have high heteroplasmic variation.

For MESA sample SRR9247860, we found that having both iterations of heteroplasmy calls is useful. In the second iteration position 3666 was annotated as a multiallelic site but only listed one allele A > C (23%). However, when we evaluated at the first iteration variants, position 3666 had two alternative alleles for the G rCRS ref allele G > A (75.9%) and G > C (23.5%) with a combined frequency of 99.4%. To get up to 100% frequency, the G allele must be present at <1%, making this a triallelic site. Our pipeline outputs the full list of variant calls to aid in understanding multiallelic variant sites. It is important to note that a direct ref/alt allele comparison of first and second iteration heteroplasmy variants will miss much of the features in heteroplasmy identification.

To further validate our approach of calling heteroplasmy, after remapping to each samples unique reference mtDNA sequence, we called heteroplasmy on the same 30 simulated datasets described above. At low coverage, running the second iteration of Mutect2 variant calling was the most accurate at identifying the known heteroplasmic sites in our simulated dataset (Table 2). The second iteration identified fewer false positives at 1.7 sites compared to 1.9 sites using just the first iteration of Mutect2. The second iteration of Mutect2 only detected true positive variants at 2000x coverage making it considerably better than other methods at variant calling in our simulated data. We use the second iteration Mutect2 variant calls for further analyses.

**Table 2:** Average Number of Heteroplasmic Sites Identified Across 30 Simulated Data

|  | All Identified sites | True positives | False positives |
| --- | --- | --- | --- |
| **100x coverage** |  |  |  |
| 1<sup>st</sup> iteration Mutect2 | 43.27 | 41.80 | 1.91 |
| 2<sup>nd</sup> iteration Mutect2 | 42.93 | 41.80 | 1.70 |
| **2000x coverage** |  |  |  |
| 1<sup>st</sup> iteration Mutect2 | 43.03 | 42.93 | 1.00 |
| 2<sup>nd</sup> iteration Mutect2 | 43.00 | 43.00 | 0 |

### 3.9 Heteroplasmy Association with Age and mtDNA-CN

We investigated the association between the number of SNV heteroplasmic sites and age using a negative binomial generalized linear model, adjusted for sex, collection center, and race. SNV heteroplasmic site counts were from the second iteration of variant calling. Due to the variability of variant calls at chrM homopolymer regions, we counted non-homopolymer and homopolymer heteroplasmic sites separately. As expected, the number of heteroplasmies increased with age and this effect was stronger for the non-homopolymer variant site count (Figure 7 and Table 3). The beta estimates were similar for ARIC and MESA, indicating that in both cohorts, we observe a similar age effect on heteroplasmy despite differences in the age distributions of the cohorts (ARIC=45-74, MESA=44-84). Additionally, the effect of age on the number of heteroplasmies was 3x larger for non-homopolymer sites but there is still a significant association with homopolymer sites. This suggests that for homopolymer sites there may be a real, biological signal with a reduced effect size due to the inclusion of false-positive calls. We investigated the association of SNV heteroplasmic count with sex-adjusted mtDNA-CN. While we observed a general trend that as mtDNA-CN decreases, heteroplasmy increases (Table 4), as previously shown[32], there was extensive heterogeneity of the results. The lack of consistency with respect to effect size in homopolymer and non-homopolymer sites between the 2 cohorts make it challenging to draw clear conclusions of the results.

**Figure 7:**
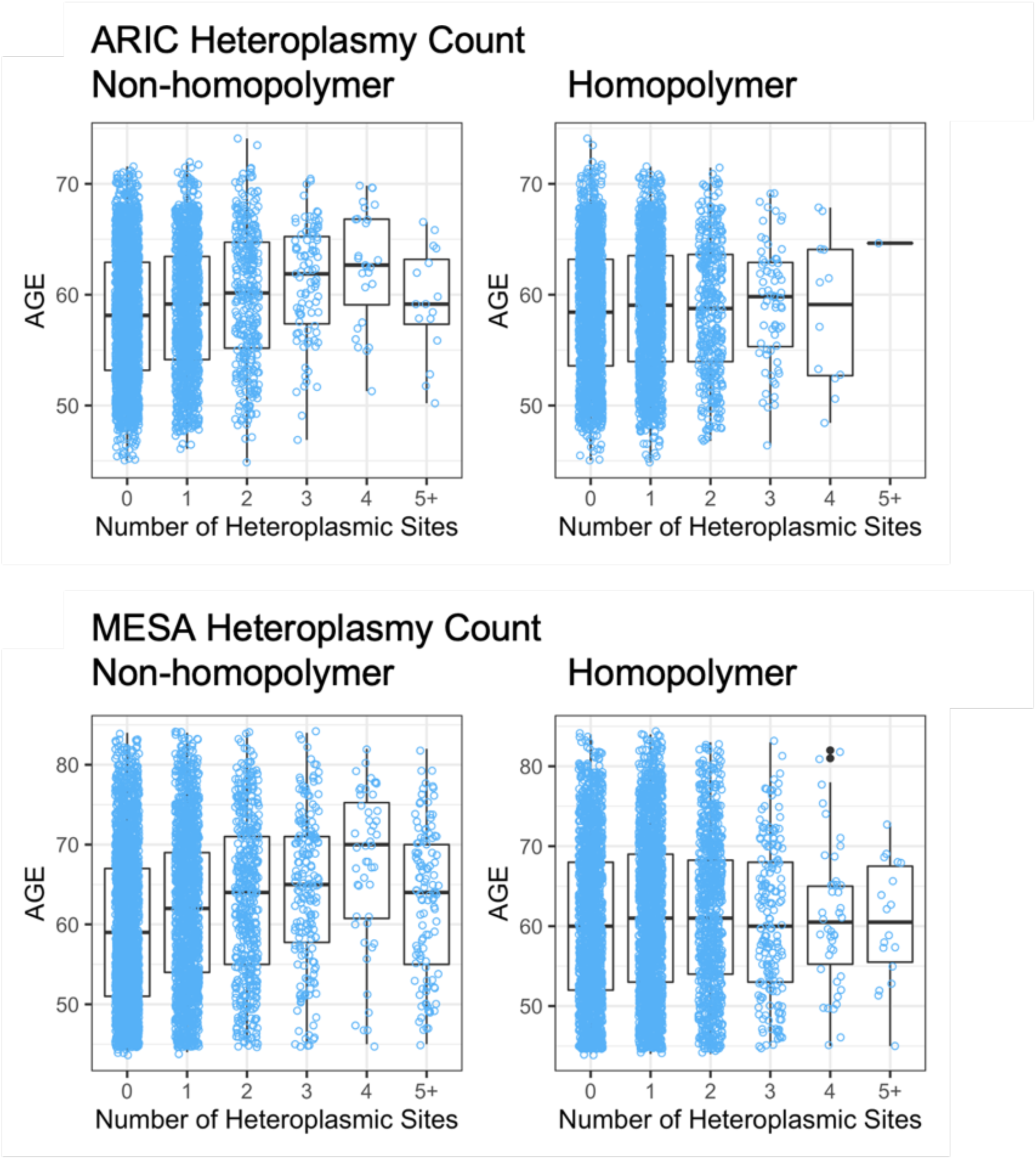
Heteroplasmy by Age Boxplots showing the age distribution of samples categorized by the heteroplasmic site count. X axis corresponds to the count of heteroplasmic sites in a sample. Category ‘“5.” contains all samples with 5 or more heteroplasmic sites.

**Table 3:** Association of Heteroplasmic Site Count with Age Age association with the presence of at least one heteroplasmic site. Heteroplasmic sites are counted by location: in a homopolymer region or in a non-homopolymer region for each sample in each cohort.

| Age Association with Heteroplasmy |  |  |  |  |
| --- | --- | --- | --- | --- |
| Location | Estimate | Std. Error | t value | p value |
| ARIC |  |  |  |  |
| Non-homopolymer | 0.015 | 0.003 | 5.441 | 5.66E-08 |
| Homopolymer | 0.005 | 0.003 | 1.681 | 0.0929 |
| MESA |  |  |  |  |
| Non-homopolymer | 0.014 | 0.001 | 9.332 | 1.58E-20 |
| Homopolymer | 0.004 | 0.002 | 2.378 | 0.0174 |

**Table 4:** Association of Heteroplasmy site count with mtDNA-CN mtDNA-CN association with the presence of at least one heteroplasmic site. Heteroplasmic sites were counted by location: in a homopolymer region or in a non-homopolymer region for each sample in each cohort. mtDNA-CN was measured using the sex-adjusted metric.

| mtDNA-CN and associations with Heteroplasmy |  |  |  |  |
| --- | --- | --- | --- | --- |
| Location | Estimate | Std. Error | t value | p value |
| ARIC |  |  |  |  |
| Non-homopolymer | -0.00016 | 0.00016 | -0.94459 | 0.345 |
| Homopolymer | -0.00062 | 0.00016 | -3.75267 | 0.000178 |
| MESA |  |  |  |  |
| Non-homopolymer | -0.00113 | 0.00019 | -5.96937 | 2.56E-09 |
| Homopolymer | -0.00032 | 0.00019 | -1.66788 | 0.0954 |

### 3.10 INDELs

INDELs were not as consistently identified in the chrM, with high variability between the number of INDELs called by Mutect2 and Mutserve. In MESA, INDEL site count in Mutect2 and Mutserve had an adjusted R-squared of 0.19 for heteroplasmy and 0.06 for homoplasmy. In ARIC, INDEL site count at a 3% VAF threshold had an adjusted R-squared value of 0.17 and 0.05 for heteroplasmic and homoplasmic INDELs, respectively. We further examined the INDELs identified by Mutect2 since Mutserve is not designed for INDEL identification. In ARIC and MESA, respectively, of the >10,000 and >17,000 total INDELs identified occurred at only 119 and 195 unique sites, with 60% in ARIC and 43% in MESA found within 150 bases (the length of an Illumina sequence read) of one of the 9 homopolymer regions on chrM rCRS sequence (Figure 8). These regions are prone to PCR, sequencing, and mapping error, and thus at this time we are not able to confidently evaluate them.

**Figure 8.**
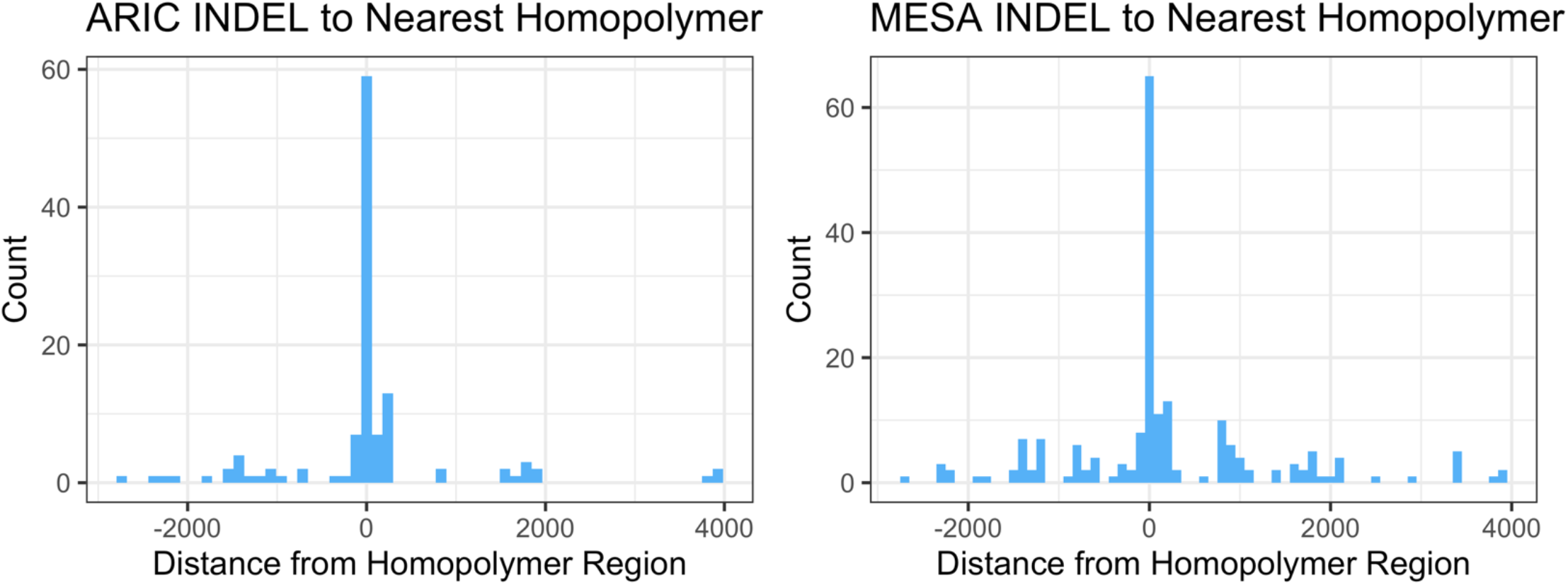
Location of INDEL variants relative to homopolymer regions The x-axis is the distance from an INDEL to its nearest homopolymer region. INDELs that directly overlap a region have a distance of 0.

## 4 Discussion

Here we present a bioinformatics pipeline to analyze mtDNA-CN and heteroplasmy from WGS data. We optimize our pipeline to run quickly and efficiently on 10,000s of samples, a necessity for large scale genomics studies. We built into our pipeline methods to recover chrM reads at the ends of chrM and at other typically low coverage regions by remapping unmapped reads to a circularized chrM sequence and by remapping reads from NUMTs on chr1 and chr17. These sequences are chrM read “sinks” and with 150 bp paired-end data, we are able to appropriately map them to chrM. We investigated different calculations for mtDNA-CN, and found that there was no significant difference between them. We demonstrate that we are able to detect true population-level SNV variants at 2000x chrM coverage, as shown by our high overlap with variants in the gnomAD database. The most novel aspect of our pipeline is the second iteration variant identification, which calls variants using a sample’s unique chrM sequence as the reference. We validate this technique’s ability to remove false positive variants and accurately call true positives using simulated data. To date, no other heteroplasmy software takes this approach. Our method outperforms existing software in assessing intra-individual heteroplasmic load by identifying heteroplasmic variants against an individual’s unique chrM reference sequence rather than a standard reference. We found that homopolymer regions in chrM give most of the variability in heteroplasmy calls and but still have an association with increasing age although weaker than heteroplasmic sites in non-homopolymer regions. Within our output files, we provide annotated VCFs. Many of these annotations are taken directly from Mutect2 and others are described on our GitHub: https://github.com/danarking1/HP. All-in-all, we demonstrate that our pipeline has increased accuracy and precision in mtDNA variant calling over other existing pipelines and we provide recommendations on how to interpret the data outputs.

We built our pipeline to run on human data already mapped to hg19 or hg38 reference genomes. However, this pipeline could easily be adopted for other organisms with known mitochondrial variation. There are other interesting facets of mitochondrial genetics that can also be addressed through our pipeline. First, ARIC and MESA used slightly different methods for isolating cells for DNA extraction. It would be interesting to investigate how subtle differences in the blood cell composition could affect the heteroplasmic variants we detect. Second, the analyses presented used the rCRS but we have made RSRS an optional reference genome in our pipeline. This may affect the homoplasmic variants identified but should have no effect on second iteration of heteroplasmic variant calls. Currently our pipeline only uses WGS data but later versions may accept other inputs such as whole exome sequencing data, RNA-seq or microarray data or FASTQ format sequencing reads as an input.

There are a few limitations to our approach. The work presented here uses paired-end 150bp sequencing reads. WGS data with shorter read lengths or from different DNA sequencing platforms may perform differently in our pipeline. Variants identified in homopolymer regions and/or hypervariable regions tend to also have annotations such as strand bias and clustered event. The software identifies these regions as potential sequencing errors. Variants in these regions should be evaluated cautiously or excluded altogether, keeping in mind that many of these variants reside in the D-loop, a region with high sequence variation[33]. We leave it to the user’s discretion how to handle these variants. INDELs are prone to occur in homopolymer regions. Some optimization is still needed for INDEL variant identification to take into account these INDEL hotspots. Currently our pipeline and the heteroplasmy variant identification software are not optimized for INDEL calling. Before assessing the biological significance, it would be advantageous to validate uncertain variants with a secondary method.

It is also important to note that in addition to the haplocheck output, there are other ways to identify problematic samples in a dataset. Any samples that are outliers for mtDNA-CN or heteroplasmy count should be considered carefully. Samples with a relatively high number of haplogroup specific, hypervariable, or hotspot variants should be closely inspected. Moreover, samples with a relatively high number of NUMT annotated variants should also be examined prior to downstream analyses. As with any experimental data, it is important to consider results of data analysis within the context of the biology of the samples in question.

Our bioinformatics pipeline captures many features of mitochondrial genetics that are important in understanding the contribution of the mtDNA to disease. As with any variant identification software we recommend variants of high interest be validated with a secondary method. However we present a framework for accelerating the analysis of mitochondrial genetics.

## Data Availability

No new sequencing data were created for this study. Sequencing data used in this study is available through dbGap (https://www.ncbi.nlm.nih.gov/gap/): NHLBI TOPMed - NHGRI CCDG: Atherosclerosis Risk in Communities (ARIC) (phs001211.v4.p3); NHLBI TOPMed: MESA and MESA Family AA-CAC (phs001416.v2.p1). All data are available from the corresponding author upon reasonable request.

## Data Availability

No new sequencing data were created for this study. Sequencing data used in this study is available through dbGap (https://www.ncbi.nlm.nih.gov/gap/): NHLBI TOPMed -NHGRI CCDG: Atherosclerosis Risk in Communities (ARIC) (phs001211.v4.p3); NHLBI TOPMed: MESA and MESA Family AA-CAC (phs001416.v2.p1). All data are available from the corresponding author upon reasonable request.

## Acknowledgements

TOPMed Acknowledgements

Molecular data for the Trans-Omics in Precision Medicine (TOPMed) program was supported by the National Heart, Lung and Blood Institute (NHLBI). Genome Sequencing for “NHLBI TOPMed: NHGRI CCDG: Atherosclerosis Risk in Communities (ARIC)” (phs001211.v4.p3) was performed at Baylor (3U54HG003273-12S2; HHSN268201500015C). Genome Sequencing for “NHLBI TOPMed: MESA and MESA Family AA-CAC” (phs001416.v2.p1) was performed at Broad Genomics (3U54HG003067-13S1; HHSN268201600034I). Core support including centralized genomic read mapping and genotype calling, along with variant quality metrics and filtering were provided by the TOPMed Informatics Research Center (3R01HL-117626-02S1; contract HHSN268201800002I). Core support including phenotype harmonization, data management, sample-identity QC, and general program coordination were provided by the TOPMed Data Coordinating Center (R01HL-120393; U01HL-120393; contract HHSN268201800001I). We gratefully acknowledge the studies and participants who provided biological samples and data for TOPMed.

ARIC Acknowledgements

Whole genome sequencing (WGS) for the Trans-Omics in Precision Medicine (TOPMed) program was supported by the National Heart, Lung and Blood Institute (NHLBI). WGS for “NHLBI TOPMed: Atherosclerosis Risk in Communities (ARIC)” (phs001211) was performed at the Baylor College of Medicine Human Genome Sequencing Center (HHSN268201500015C and 3U54HG003273-12S2) and the Broad Institute for MIT and Harvard (3R01HL092577-06S1). Centralized read mapping and genotype calling, along with variant quality metrics and filtering were provided by the TOPMed Informatics Research Center (3R01HL-117626-02S1).

Phenotype harmonization, data management, sample-identity QC, and general study coordination, were provided by the TOPMed Data Coordinating Center (3R01HL-120393-02S1). We gratefully acknowledge the studies and participants who provided biological samples and data for TOPMed. The Genome Sequencing Program (GSP) was funded by the National Human Genome Research Institute (NHGRI), the National Heart, Lung, and Blood Institute (NHLBI), and the National Eye Institute (NEI). The GSP Coordinating Center (U24 HG008956) contributed to crossprogram scientific initiatives and provided logistical and general study coordination. The Centers for Common Disease Genomics (CCDG) program was supported by NHGRI and NHLBI, and whole genome sequencing was performed at the Baylor College of Medicine Human Genome Sequencing Center (UM1 HG008898 and R01HL059367).

MESA Acknowledgements

This work was supported by grants from the National Institutes of Health [R01HL131573; R01HL144569] Whole genome sequencing (WGS) for the Trans-Omics in Precision Medicine (TOPMed) program was supported by the National Heart, Lung and Blood Institute (NHLBI). WGS for “NHLBI TOPMed: Multi-Ethnic Study of Atherosclerosis (MESA)” (phs001416.v2.p1) was performed at the Broad Institute of MIT and Harvard (3U54HG003067-13S1). Centralized read mapping and genotype calling, along with variant quality metrics and filtering were provided by the TOPMed Informatics Research Center (3R01HL-117626-02S1; contract HHSN268201800002I). Phenotype harmonization, data management, sample-identity QC, and general study coordination, were provided by the TOPMed Data Coordinating Center (3R01HL-120393-02S1; contract HHSN268201800001I). The MESA project is conducted and supported by the National Heart, Lung, and Blood Institute (NHLBI) in collaboration with MESA investigators. Support for MESA is provided by contracts 75N92020D00001, HHSN268201500003I, N01-HC-95159, 75N92020D00005, N01-HC-95160, 75N92020D00002, N01-HC-95161, 75N92020D00003, N01-HC-95162, 75N92020D00006, N01-HC-95163, 75N92020D00004, N01-HC-95164, 75N92020D00007, N01-HC-95165, N01-HC-95166, N01-HC-95167, N01-HC-95168, N01-HC-95169, UL1-TR-000040, UL1-TR-001079, UL1-TR-001420. Also supported in part by the National Center for Advancing Translational Sciences, CTSI grant UL1TR001881, and the National Institute of Diabetes and Digestive and Kidney Disease Diabetes Research Center (DRC) grant DK063491 to the Southern California Diabetes Endocrinology Research Center.

