## Supplemental Figures for "A Bioinformatics Pipeline for Estimating Mitochondria DNA Copy Number and Heteroplasmy Levels from Whole Genome Sequencing Data"

**Affiliations:**
1 McKusick-Nathans Institute, Department of Genetic Medicine, Johns Hopkins University School of Medicine, Baltimore, MD, USA
2 Department of Biomedical Engineering, Johns Hopkins University, Baltimore, MD, USA
3 Human Genetics Center, Department of Epidemiology, Human Genetics, and Environmental Sciences, School of Public Health, The University of Texas Health Science Center at Houston, Houston, TX, USA
4 Human Genome Sequencing Center, Baylor College of Medicine, Houston, TX, USA
5 The Institute for Translational Genomics and Population Sciences, Department of Pediatrics, The Lundquist Institute for Biomedical Innovation at Harbor-UCLA Medical Center, Torrance, CA, USA
6 Center for Public Health Genomics, University of Virginia, Charlottesville, VA, USA
7 Department of Laboratory Medicine and Pathology, University of Minnesota Medical School, Minneapolis, MN, USA
8 Evans Department of Medicine and the Whitaker Cardiovascular Institute, Boston University School of Medicine, Boston, MA, USA
9 Framingham Heart Study, Boston University School of Medicine, Boston, MA, USA.
10 Department of Biostatistics, Boston University School of Public Health, Boston, MA, USA

*co-first authors

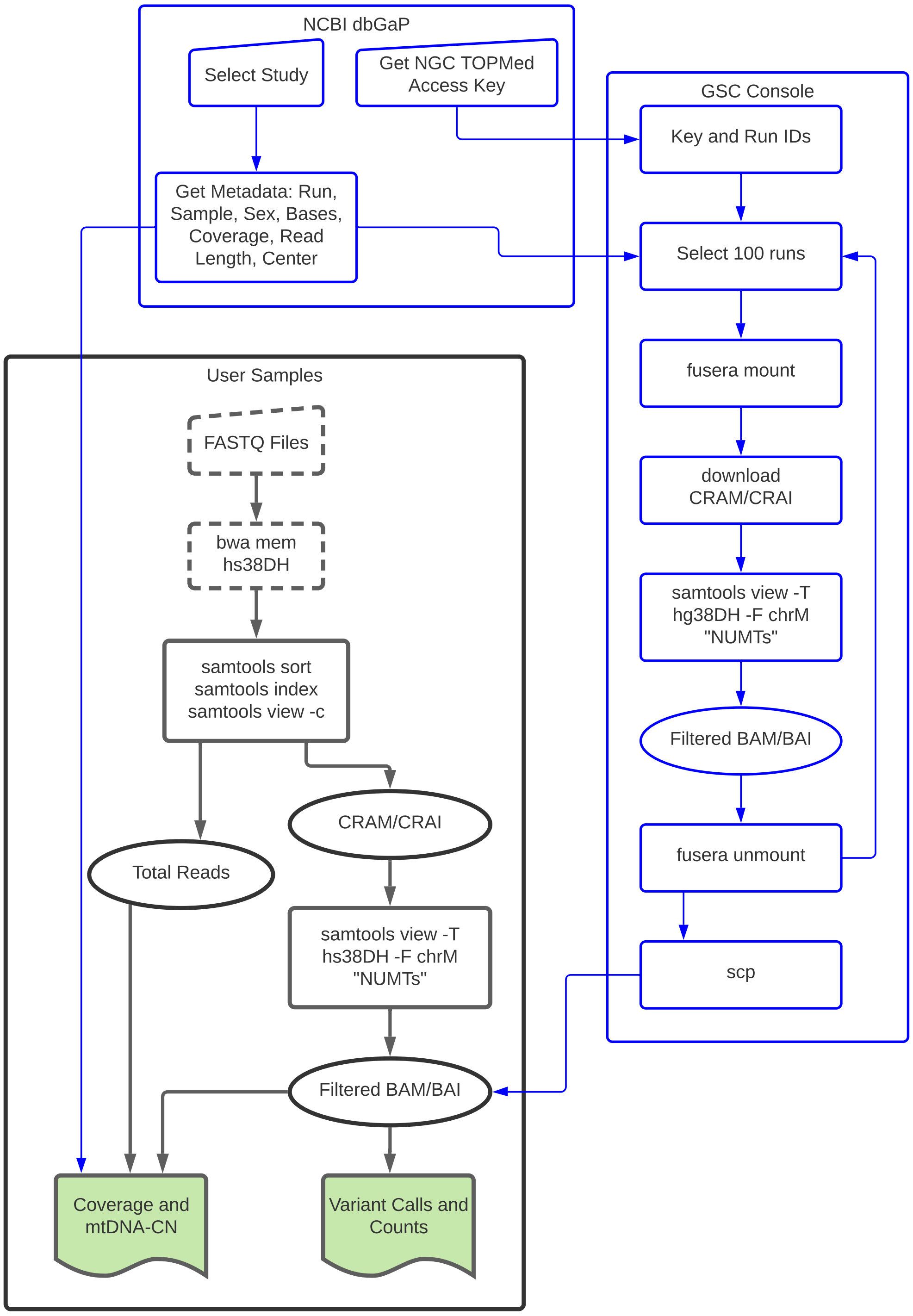


**Supplementary Figure 1**
Flowchart for accessing TOPMed data or user specific input data. Blue boxes were performed on Google Cloud Services. Dashed boxes are steps performed by user prior to running the pipeline.

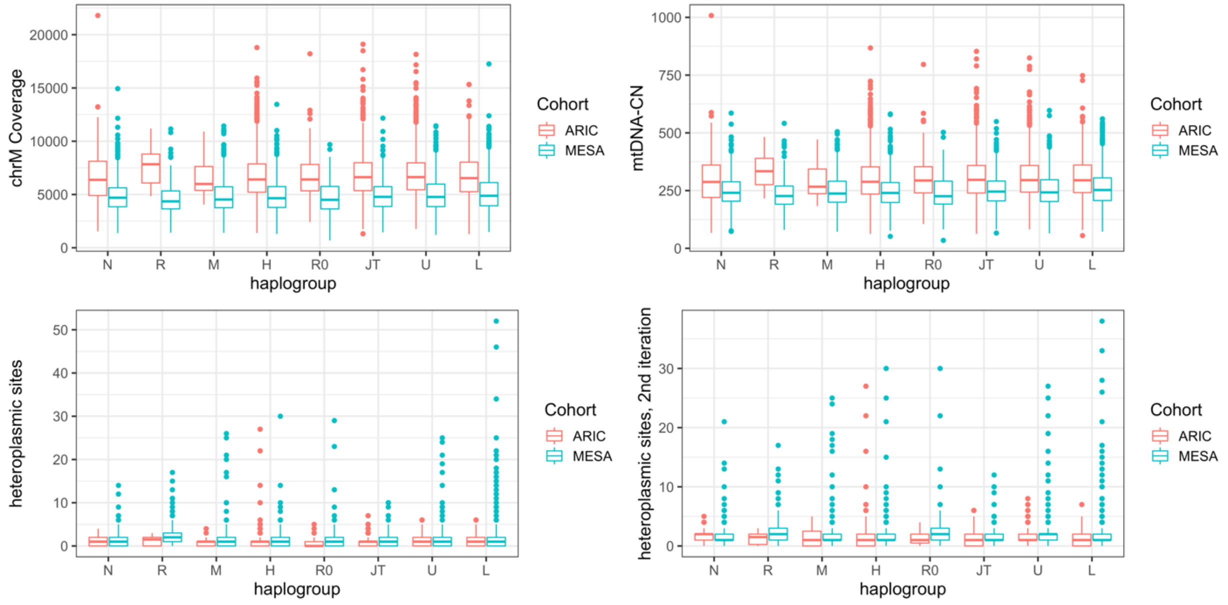


**Supplementary Figure 2**
Haplogroups are grouped accordingly: H is H only; N contains N, Y, A, S, I, W, X; R contains R1-9, B, P F; M contatins M, C, Z, E, D, G, Q; R0 contains R0, HV, V; U contains U, K; JT contains J, T; L contains L0-6. No significant differences were observed between haplogroups for chrM coverage or mtDNA-CN. For heteroplasmic SNV site count compared to group H, groups R0 and U in ARIC differed (p values 0.02 and 0.0002) and in MESA groups R, M, R0, U and L were significantly different (p values 1.24e-12, 0.01, 0.001, 0.009, 0.0006). For the second iteration of heteroplasmic site count, in ARIC, groups N, JT, U, and L were significantly different (p values 0.0005, 0.007, 0.007, and 0.047) and in MESA groups R, M, R0, and U were significantly different (p values 2.37e-11, 0.01, 0.0003, and 0.003).

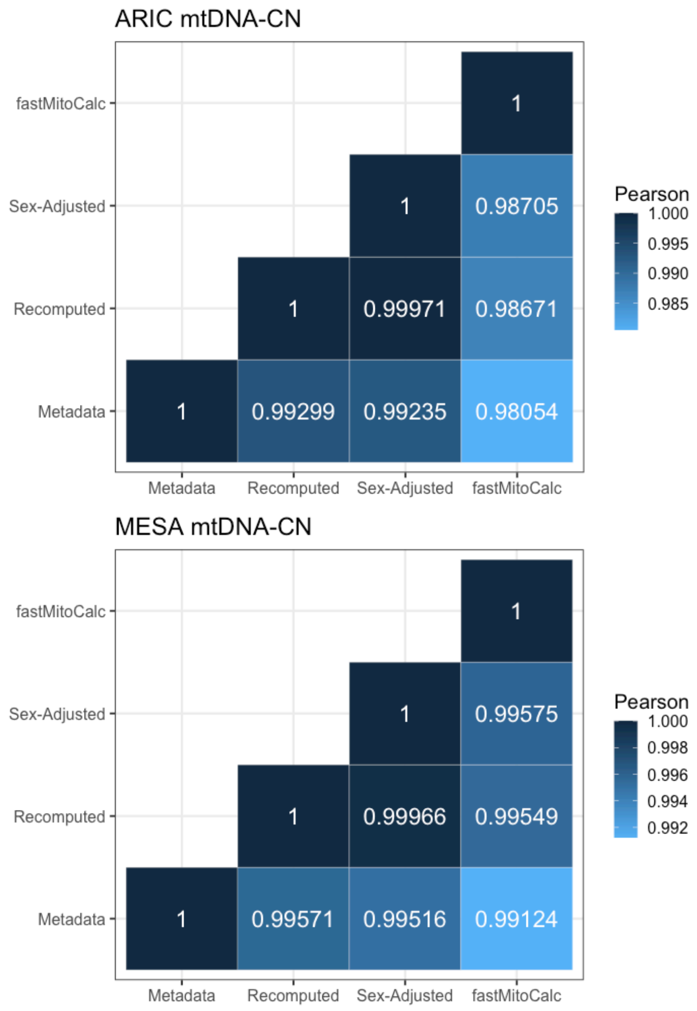


**Supplementary Figure 3:** Correlation between mtDNA-CN metrics
Pairwise correlation between the different mtDNA-CN calculated metrics is plotted in the matrix for ARIC and MESA cohorts. The Pearson correlation coefficient is written inside the square for each pairwise comparison.

| mtDNA-CN Metric | ARIC Age | ARIC Sex | mtDNA-CN PRS | MESA Age | MESA Sex | Mean Rank | Kendalls W p value |
|---|---|---|---|---|---|---|---|
| sex adjusted | 2 | 2 | 3 | 3 | 1 | 2.2 | 0.56 |
| recomputed | 1 | 3 | 2 | 2 | 3 | 2.2 | |
| metadata | 3 | 4 | 4 | 1 | 4 | 3.2 | |
| fastMitoCalc | 4 | 1 | 1 | 4 | 2 | 2.4 | |

**Supplementary Figure 4:** Rankings of mtDNA-CN Strength of Associations. Significance of the ranking by cohort is shown by the Kendall's W p value.

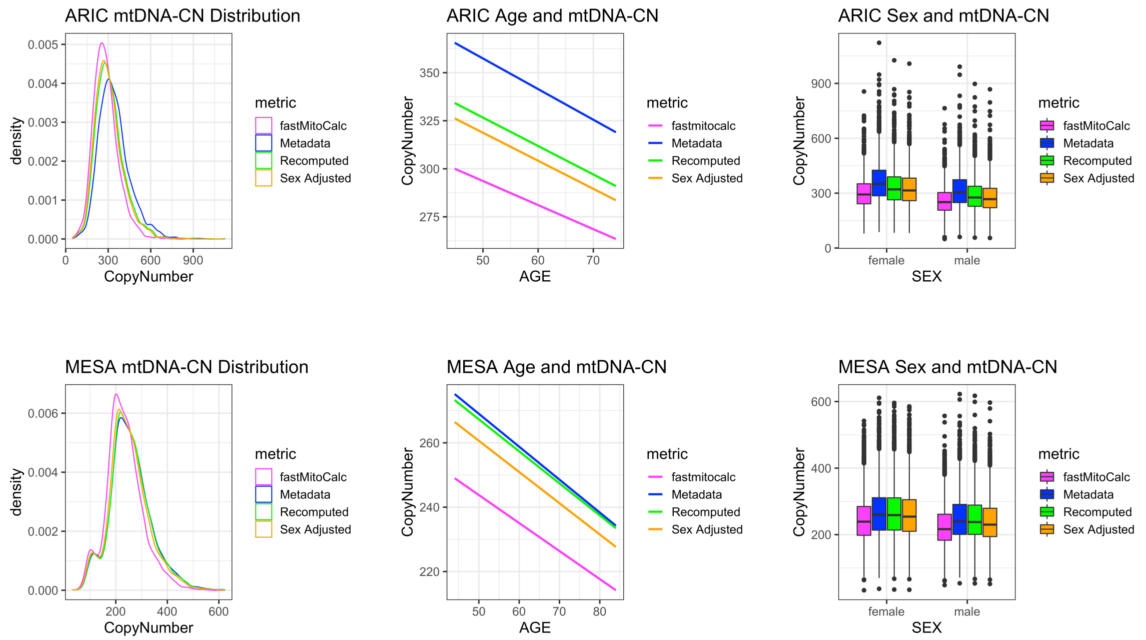


**Supplementary Figure 5**

Graphs of mtDNA-CN metrics in ARIC (top row) and MESA (bottom row). From left to right: density plot of mtDNA-CN distribution, graph depicting the relationship between sample age (x-axis) and mtDNA-CN (y-axis), and boxplots of mtDNA-CN by sex.

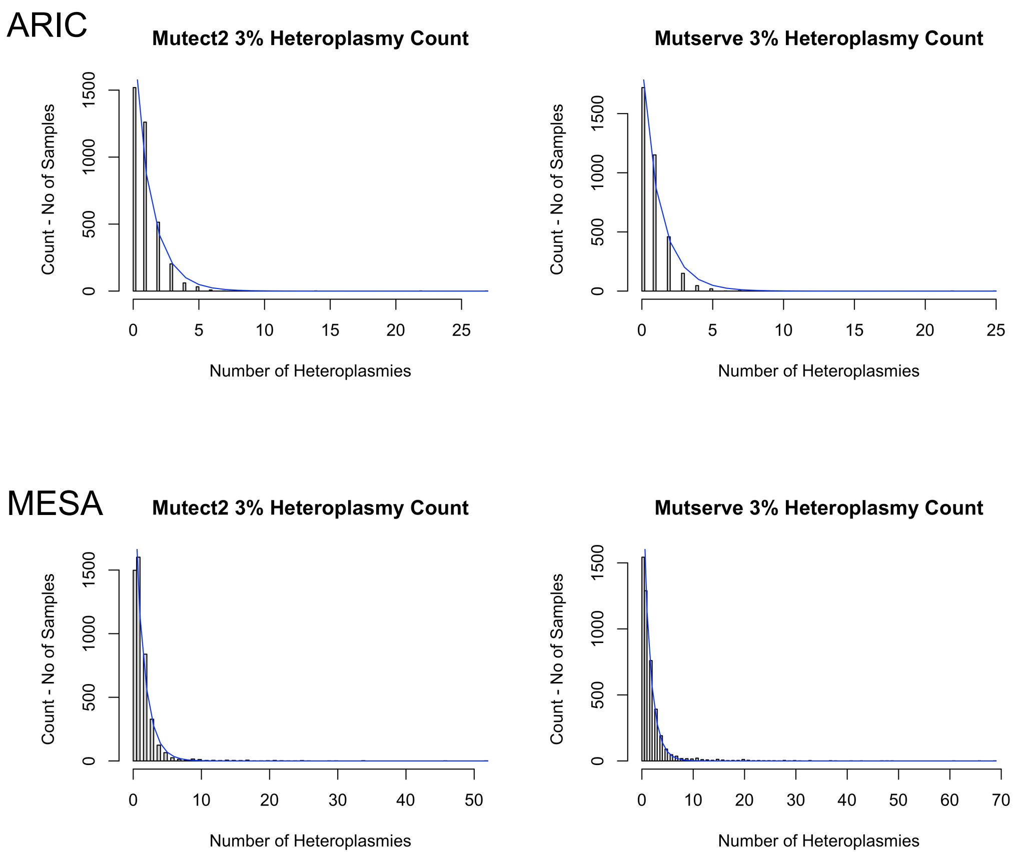


**Supplementary Figure 6**
Distribution of heteroplasmic site count from Mutect2 and Mutserve identified variants.

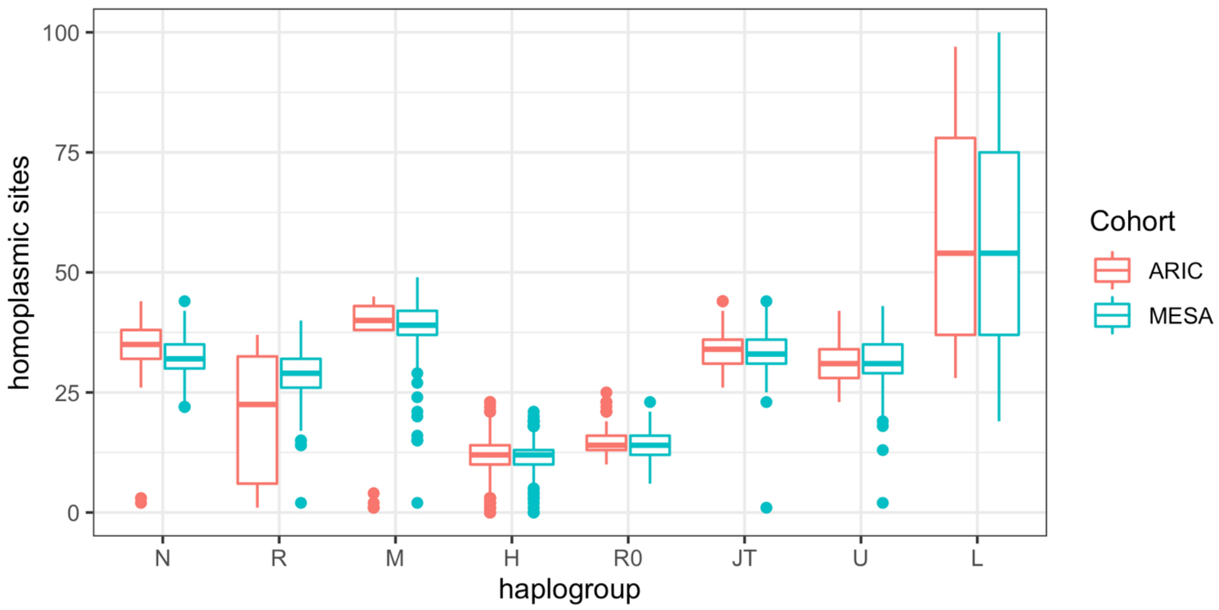


**Supplementary Figure 7**
Boxplots showing the distribution of chrM sequencing coverage (upper left), mtDNA-CN sex adjusted metric (upper right), heteroplasmic site count from the first iteration variant calling (lower left), and heteroplasmic site count from the second iteration variant calling (lower right). Samples are grouped by haplogroup as defined in Supplementary Figure 2 figure legend.

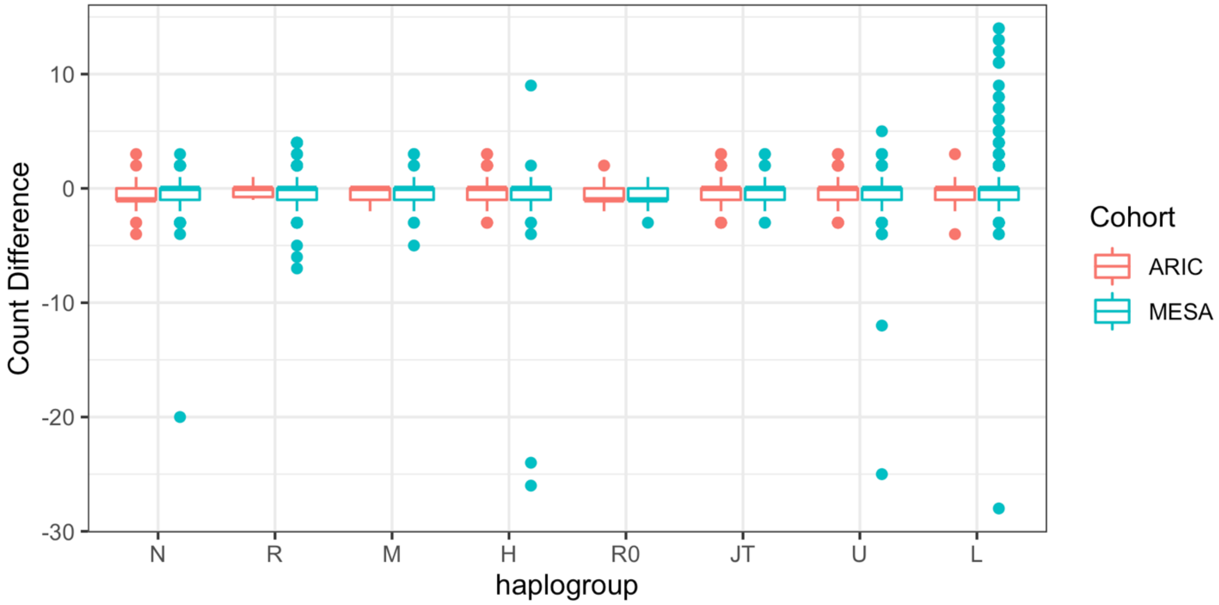


**Supplementary Figure 8**
Count Difference is defined as the first iteration SNV heteroplasmic site count minus the second iteration heteroplasmic site count. Samples are grouped by haplogroup as defined in Supplementary Figure 2 figure legend.
